# Correlation between religiosity and family functioning among secondary school students in high-risk residing areas and factors associated with substance use

**DOI:** 10.1101/2024.07.21.24310785

**Authors:** Noor Adnin binti Ab Aziz, Suzaily Wahab, Rosnah binti Sutan, Muhammad Adib Baharom, Amirul Danial Azmi, Siti Azirah binti Asmai

## Abstract

**Introduction:** Substance use in adolescents poses a complex societal challenge that undermines nation-building and socioeconomic growth. Religiosity refers to a person’s religious beliefs, habits, and involvement in religious activities. Family functioning refers to the overall health and operation of a family unit, which includes communication, emotional bonding, support, roles, and behavioral control. Both aspects play a significant impact in determining substance use in adolescents. This study is to assess the correlation between religiosity, and family functioning, and to determine factors associated with substance use among adolescents in secondary schools in high-risk areas.

**Methods:** A cross-sectional study was conducted among 312 adolescents from selected secondary schools in substance use hotspot areas in Northern Malaysia. Alcohol, Smoking, and Substance Involvement Screening Tool-Lite (ASSIST-Lite), Family Adaptation and Cohesion Scale version IV (FACES-IV), and Hatta Islamic Religiosity Scale (HIRS) were used as instruments.

**Results:** The prevalence of substance use among adolescents was 9.6%(n=30). Most of the users used a single substance (76.7%; n=23) and only (23.3%; n=7) used multiple substances. The mean age was 14.13 years (SD:0.67), and the majority were Malays (99.0%; n=309) with a background in Muslim religion. Adolescent substance use was significantly associated with gender (16.3% in males and 6.3% in females) and having a recent family history of substance use (16.8%). A negative correlation was found between substance use and family functioning; balanced flexibility (rs=-0.12; P=<0.05), family communication (rs=-0.12; P=<0.05), and family satisfaction (rs=-0.15; P=<0.01). There was a positive correlation between substance use and chaotic family (rs=0.12; P=<0.05). Regression analysis reveals that only male adolescents and a recent family history of substance use were significant predictors of substance use. Family satisfaction was the only significant protective factor. There was no significant association between substance use and religiosity (rs=-0.01; P= 0.83).

**Conclusions:** These findings can assist policymakers, healthcare professionals and schools develop interventions to reduce substance use, especially in high-risk communities, and increase adolescents’ well-being in general.

## Introduction

Substance or drug use is a worldwide problem and the prevalence of substance use among adolescents reveals a rising trend of use [1,2]. World Drug Report 2023 revealed in 2021, one out of every seventeen people aged 15 to 64 in the world have used a drug within the previous year [3]. Substance or drug use refers to the consumption or intake of psychoactive substances, including tobacco, alcohol, prescription medications, and illicit drugs. Tobacco and alcohol are examples of licit psychoactive substances, while illicit substances include amphetamines, heroin, cocaine, marijuana, and lysergic diethylamide (LSD). It has a substantial impact on public services, the criminal justice system, and health care services in the long term [4]. In Malaysia, the National Anti-Drugs Agency Ministry of Home Affairs reports that from 2016 to 2020, the number of substance and drug use detected by the state and among adolescents exhibited an upward trend as well. Synthetic drugs appear to be more commonly used than organic drugs. The most used illicit drugs in Malaysia include methamphetamine (crystalline), amphetamine-type stimulants, heroin and morphine, methamphetamine (pills), and cannabis.

Religiosity, defined as the extent and intensity of a person’s religious beliefs, practices, and experiences, varies greatly between cultures [5]. It has a substantial impact on adolescents, their behaviors, and lifestyle choices, including substance use. In Western countries, religion frequently stresses personal belief and individualized spirituality, with a high degree of secularism and a wide range of religious affiliations. Numerous studies have found that religiosity serves as a protective factor for substance use [6–9]. In ASEAN countries, religiosity is more communal and profoundly incorporated into daily life, with state-endorsed religions such as Islam in Malaysia [10]. The majority of Malays in Malaysia practice Islam as their religion. They are taught fundamental knowledge and practice about Islam since primary school. A few local research found that strong religious views were protective of substance use among Malaysian youths [11,12]. Given this prior information about the students and their devotion to Islam, as well as the lack of research on the relationship between religiosity and substance use among adolescents, the fact that many Malay adolescents continue to use substances is alarming. This study aims to assess religiosity among adolescents in terms of knowledge and practice following Islamic teachings and the correlation with substance use.

Previous research has shown that a person’s physical and psychological development is profoundly affected by their family. Family functioning is defined as the quality and health of interactions and relationships within a family unit. A family’s primary functions include instilling beliefs, values, and acceptable behavior in society, particularly during childhood [13]. Many previous literatures reported poor family functioning as s significant factor that contributed to substance use [14–16]. Family functioning in this study refers to family cohesion and adaptability. Family cohesion is the emotional connection of family members, an instance in which all members of the family are accountable for one another, whereas adaptability refers to the extent to which a family structure is adaptable to changes.

In Malaysia, the Malaysian Anti-Drug Agency (AADK) identified 155 drug-related hotspots in 2020. The northern region of Malaysia, which comprises a few states in association with the Thai border, is recognized as a hotspot for substance usage [17]. Adolescents in hotspot areas may have a greater predisposition for substance use than those in non-hotspot areas. Adolescence are the group vulnerable to drug use and addiction as they have a high tendency toward risking new experiences, and novelty, are prone to peer pressure, and have poor self-esteem [18]. This issue may start at 12 to 17 years and may crown between the ages of 18 and 25 years [19]. The earlier a person begins engaging in substance use, the higher their chance of developing substance-use disorder. The use of other illicit substances is often preceded by using-licit substances, such as tobacco and alcohol [20]. More harmful consequences on health and psychosocial wellness in adolescents compared to adults as it often lasts longer into their lives because of joblessness, monetary reliance, and deficient social affairs. 10,600 adolescents in Malaysia have used drugs at least once in their lives [21].

In keeping with the numerous factors that contribute to adolescent substance use and the lack of studies done locally involving adolescents in the hotspot areas, our study intended to evaluate the relationship between substance use, religiosity, and family functioning among adolescents.

## Methodology

### Research design, study area, and data collection

A cross-sectional study was conducted in selected secondary schools from the Northern Malaysia region. The data was obtained using stratified random sampling from December 1st to 15th, 2023. The class teachers and students were informed about the study’s methods, objectives, participation benefits, and risks, affirmed anonymity, and the ability to leave at any point. Those who obtained consent from both their parents and them were asked to fill out questionnaires using Google Forms, an online survey platform. Duplicate responses were removed during data cleaning. This study was approved by the Institution’s research ethics committee (UKM PPI/111/8/JEP-2023-076) and granted permission from the Education Planning and Research Division, Ministry of Education Malaysia.

The sample size was calculated using the formula N = Z^2^P(1-P)/d2 (Kish L 1969). Since the previous study’s prevalence was 17.2% [16], with a precision of 0.05, the sample size needed was 215 respondents. After accounting for an expected 20% of non-respondents, 44 more respondents were added, resulting in a total sample size of 263. The inclusion requirements included secondary school students aged 13-16 years old, students with a background in Muslim Religion, who can understand and write Malay or English, and who have parental or carer agreements. The study excluded 17-year-olds since they are exam group students in Form 5.

### Study instruments

All respondents were assigned a sociodemographic questionnaire, the Hatta Islamic Religiosity Scale (HIRS), the Family Adaptation and Cohesion Scale version IV (FACES-IV), and the Alcohol, Smoking, and Substance Involvement Screening Tool-Lite (ASSIST-Lite). All questionnaires were translated and validated in the Malay language. Permission to use all the relevant questionnaires was obtained from the original authors.

### Alcohol, Smoking, And Substance Involvement Screening Tool-Lite (ASSIST-Lite)

Substance use was measured using a validated Malay version of ASSIST-Lite which is an abbreviated version of ASSIST. It is a screening tool to evaluate the substance use, the frequency of usage, and drug use by a variety of drugs of concern such as nicotine, alcohol, cannabis, amphetamine-type stimulants, sedatives, opioids, and other substances within the last 3 months. It consists of six core components with three to four items and two additional items. The substance score is calculated by summing points acquired for that substance. The internal and test-retest reliability of the Malay version of ASSIST-Lite were good, with alpha values ranging from 0.772 to 0.882 and Kappa values ranging from 0.8 to 1.

### The Hatta Islamic Religiosity Scale (HIRS96)

It is a 27-item scale to evaluate basic Islamic knowledge and practice among Muslim adults and adolescents in Malaysia. It was devised in 1996 by Mohamed Hatta Shaharom. It comprises 4 parts with 15 questions on basic Islamic knowledge following primary and lower secondary school curriculum, 10 questions on basic Islamic practice by the obligation of practicing Muslim based on the Holy Quran and al-Hadith, 1 question on degree of Qur’an reading, and 1 question evaluate respondent encouraging good and forbidding bad habit, which is competency, obligatory on all Muslims. For each component, a Likert score is used. This is a valid and reliable tool. It has high Inter-rater reliability for total HIRS score (0.90). The higher the score, the higher the religiosity [22].

### Family Adaptation And Cohesion Scale Version IV (FACES-IV)

It was used to assess components of family functioning, which were conceptualized using the Circumplex Model. It is a self-rated test that includes 42 items for balanced and unbalanced scales, 10 items for family communication, and 10 items for family satisfaction. FACES-IV’s reliability ranges from 0.77 to 0.89 for all six domains. It also has a high discriminant validity of 0.84-0.99 across all its domains. It includes three aspects of family behavior: cohesion, adaptability, and communication. Cohesion is the emotional bond among family members. Flexibility refers to the degree of change in family leadership, relational roles, and rules. Communication empowers a family to modulate its level of cohesion or flexibility. This study used the Malay-translated questionnaire which was pre-tested among 50 respondents. The Cronbach’s alpha for the translated Malay version of FACES-IV was 0.68.

### Statistical analysis

The acquired data was analyzed using the most recent version of the Statistical Package for Social Science (SPSS 29; SPSS Inc., Chicago, Illinois, United States) version 29. The relationships between study variables were analyzed using correlation tests. The statistical study used a p-value of < 0.05 to indicate significance. All categorical variables (gender, age, race, total household income per month, and father and mother’s highest education level) were presented in frequency, percentage, and Chi-square test. Descriptive analysis was used to determine the prevalence of substance use among Secondary School Students in high-risk residing areas. The Kolmogorov-Smirnov test was used to measure normality test. Binary logistic regression analysis was used to determine predicted factors with substance use among Secondary School Students in high-risk residing areas.

## Results

### Demographic characteristics

A total of 312 adolescents from selected secondary schools were enrolled in the study. Table 1 furnished an outline of the sociodemographics of the respondents. The students’ ages ranged from 13 to 16 with 66.7% (n = 208) being female and 33.3% (n = 104) being male. The students’ mean age was 14.13 (SD = 0.673). 99.7% (n = 309) of the adolescents were Malay. There were 69.9% (n=218) of the students from low-income families, with a monthly household income of less than RM2500. Most of their mothers and fathers were reported to have at least a secondary school level of education as their highest education level, with 93.3% (n= 291) and 92.6% (n= 289), respectively. 32.4% (n=101) of their family members were reported to have used a substance recently.

**Table 1.** Socio-demographic characteristics of participants & association with substance use.

| **Variables** | **Total number of participants**<br>**n=312 (%)** | **Substance Use**<br>**n= 30 (%)** | **Not use Substance**<br>**n=282 (%)** | **Chi-square test**<br>**$\chi^2$ (df)** | **P value** |
| --- | --- | --- | --- | --- | --- |
| **Gender** |  |  |  | 8.132 (1) | .004 |
| Girl | 208 (66.7) | 13 (6.3) | 195 (93.8) |  |  |
| Boy | 104 (33.3) | 17 (16.3) | 87 (83.7) |  |  |
| **Age** |  |  |  | 0.537 (1) | .464 |
| Early adolescent | 279 (89.4) | 28 (10.0) | 251 (90.0) |  |  |
| Middle adolescent | 33 (10.6) | 2 (6.0) | 31 (93.9) |  |  |
| **Race** |  |  |  | 0.322 (1) | .570 |
| Malays | 309 (99.0) | 30 (9.7) | 279 (90.3) |  |  |
| Non-Malays | 3 (1.0) | 0 | 3 (100.0) |  |  |
| **Total household income in the family** |  |  |  | 1.617 (1) | .203 |
| < RM2500.00 | 218 (69.9) | 24 (11.0) | 194 (89.0) |  |  |
| >RM2500.00 | 94 (30.1) | 6 (6.4) | 88 (93.6) |  |  |
| **Highest level of education for mother** |  |  |  | 0.565 (1) | .452 |
| Nil and primary school | 21 (6.7) | 3 (14.3) | 18 (84.7) |  |  |
| >Secondary school | 291 (93.3) | 27 (9.3) | 264 (90.7) |  |  |
| **Highest level of education for father:** |  |  |  | 0.793 (1) | .373 |
| Nil & primary school | 23 (7.4) | 1 (4.3) | 22 (95.7) |  |  |
| >Secondary school | 289 (92.6) | 29 (10.0) | 260 (90.0) |  |  |
| **Any family members recently use substance:** |  |  |  | 8.949 (1) | .003 |
| Yes | 101 (32.4) | 17 (16.8) | 84 (83.2) |  |  |
| No | 211 (67.6) | 13 (6.2) | 198 (93.8) |  |  |

Early adolescent (13,14 years old), Middle adolescent (15,16 years old)

Of the 312 secondary school students, (9.6%; n= 30) acknowledged using single or polysubstance recently according to ASSIST-Lite. Malay students (9.7%; n=30) in early adolescence (10%; n=28), male gender (16.3%; n=17), and highest education level of secondary school or higher for mother (9.3%; n=27) and father (10.0%; n=29), make up most recent substance users. Most of them were from low-income families with monthly total household income of less than RM2500 (11.0%; n = 24). In addition, (16.8%; n=17) indicated a recent family history of substance use.

Table 2 furnished that cigarettes were the most widely used current substance among students (4.8%; n = 15), followed by alcohol (2.2%; n = 7). Among the illicit substances, stimulants (1.6%, n = 5) were the most frequently used substances according to ASSIST-Lite.

**Table 2.** Prevalence of substance or drug use based on drug type.

| Type of Substance |  | n= 312 | % |
| --- | --- | --- | --- |
| Tobacco | Not Use | 297 | 95.2% |
|  | Use | 15 | 4.8% |
| Alcohol | Not Use | 305 | 97.8% |
|  | Use | 7 | 2.2% |
| Amphetamine-type stimulants | Not Use | 307 | 98.4% |
|  | Use | 5 | 1.6% |
| Cannabis | Not Use | 308 | 98.7% |
|  | Use | 4 | 1.3% |
| Inhalants, sedatives, hallucinogens | Not Use | 308 | 98.7% |
|  | Use | 4 | 1.3% |
| Opioids | Not Use | 309 | 99.0% |
|  | Use | 3 | 1.0% |

Whereas Table 3 outlines the prevalence of current single or polysubstance use.

**Table 3.** Prevalence of single or polysubstance Use.

| Substance use | n=30 | % |
| --- | --- | --- |
| Single Substance Use | 23 | 76.7% |
| Poly Substance Use | 7 | 23.3% |

### Descriptive data for religiosity

The religiosity profile of respondents based on the Hatta Islamic Religiosity (HIRS) Index yields a mean score of 63.35 with an SD = 8.61 indicating a higher religiosity score. Table 4 shows Islamic knowledge and practice scores were 23.04 with SD=2.88 and 35.58 with SD=7.15 respectively. Approximately more than 50% of respondents had a higher score of Islamic knowledge and practice.

**Table 4.** Religiosity profile of respondents according to HIRS.

| Descriptive Statistic | N | Minimum | Maximum | Mean | Std. Deviation |
| --- | --- | --- | --- | --- | --- |
| HIRS Total Score | 312 | 38.00 | 84.00 | 63.3526 | 8.61264 |
| HIRS Islamic Knowledge | 312 | 16.00 | 30.00 | 23.0449 | 2.87990 |
| HIRS Islamic Practice | 312 | 14.00 | 50.00 | 35.5769 | 7.15294 |

### Descriptive data for family functioning

Table 5 shows the respondents’ family profiles which were outlined in terms of the balanced scales (cohesion and flexibility), unbalanced scales (enmeshed, rigid, disengaged, and chaotic), family satisfaction, and family communication. Each scale was classified accordingly. The mean scores of the balanced measures were higher than those of the unbalanced scales among the individuals (26.00 and 19.14, respectively). The mean scores for family communication and family satisfaction were 37.37 (SD = 6.33) and 37.79 (SD = 6.18), respectively. The cohesion ratio [balanced cohesion/ (disengaged + enmeshed/2)], flexibility ratio [balanced flexibility/ (rigid + chaotic/2)] and total circumplex ratio (cohesion ratio + flexibility ratio/2) exceeded 1 indicating that their family functioning was balanced.

**Table 5.** Family functioning of respondents based on FACES IV.

| **Dimensions** | **Mean (SD)** | **Median ± IQR** | **min.-max.** |
| --- | --- | --- | --- |
| **Balanced scales:** |  |  |  |
| Cohesion | 24.79 (3.35) | 25.00 ± 4.00 | 8.0-35.0 |
| Flexibility | 27.21 (3.62) | 28.00 ± 3.00 | 8.0-35.0 |
| **Unbalanced scales:** |  |  |  |
| Disengaged | 17.40 (3.64) | 17.00 ± 5.00 | 8.0-31.0 |
| Enmeshed | 20.49 (2.53) | 21.00 ± 3.00 | 7.0-29.0 |
| Rigid | 22.95 (3.38) | 23.00 ± 4.00 | 7.0-32.0 |
| Chaotic | 15.70 (3.92) | 16.00 ± 5.00 | 7.0-27.0 |
| **Ratio scores:** |  |  |  |
| Cohesion ratio | 1.32 (0.25) | 1.31 ± 0.34 | 0.64-2.30 |
| Flexibility ratio | 1.44 (0.30) | 1.41 ± 0.31 | 0.68-3.09 |
| Total circumflex ratio | 1.38 (0.24) | 1.38 ± 0.27 | 0.72-2.38 |
| **Family scales:** |  |  |  |
| Family communication | 37.37 (6.33) | 38.00 ± 7.00 | 10.0-50.0 |
| Family satisfaction level | 37.79 (6.18) | 39.00 ± 7.00 | 10.0-50.0 |

### Correlation between religiosity and family functioning with substance use

The Kolmogorov-Smirnov test was applied statistically to determine the age distribution of the respondents. A non-parametric test (Spearman’s rho) was utilized because the respondents’ ages were not normally distributed. As reported in Table 6, there was no significant correlation found between religious knowledge and practice and substance use.

**Table 6.**
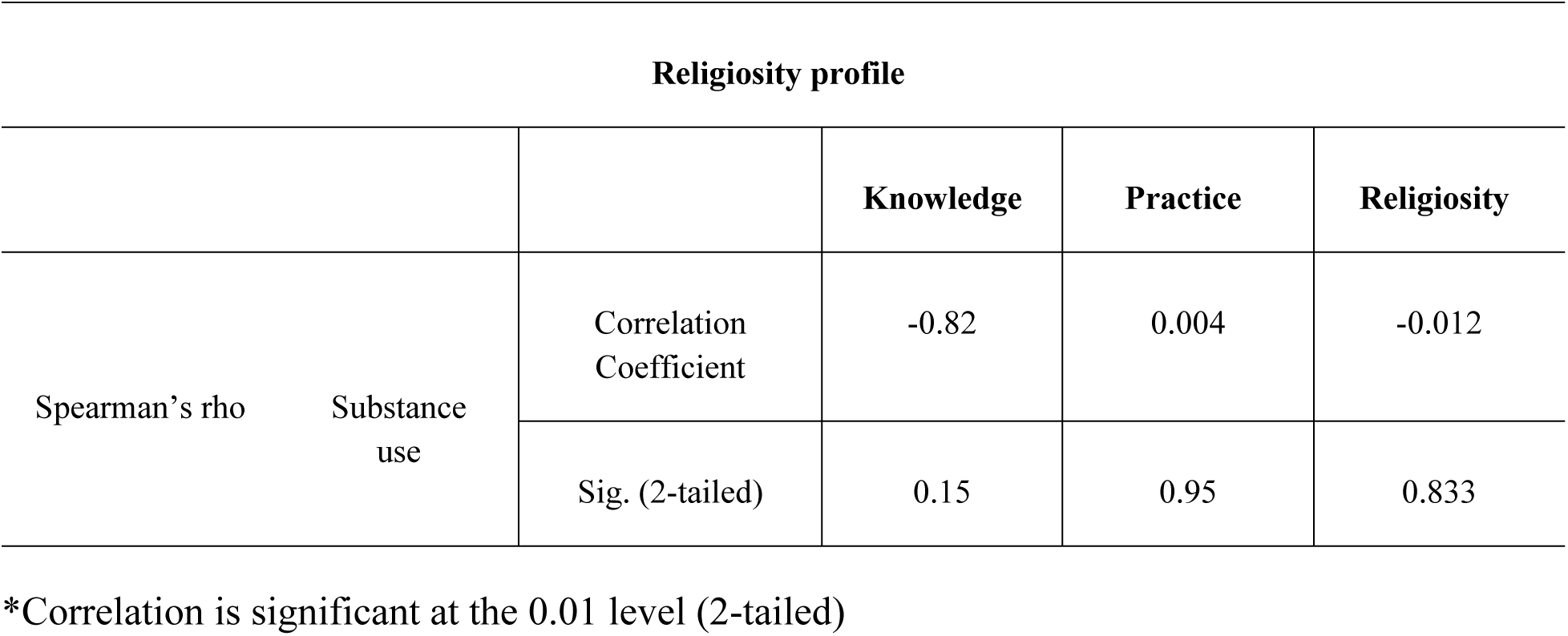
Islamic religiosity profile according to substance use Religiosity profile.

As depicted in Table 7, there is a significant negative correlation between family functioning and substance use in terms of balanced flexibility (rs=-.116; P=<.05), family communication (rs=-.116; P=<.05) and satisfaction (rs=-.153; P=<.01). On top of that, there is a significant positive correlation between chaotic family and substance use (rs=.122; P=<.05).

**Table 7.** Family functioning profile according to the substance use status.

|  |  |  | **Cohesion** | **Flexibility** | **Disengaged** | **Enmeshed** | **Rigid** | **Chaotic** | **Communication** | **Satisfaction** |
| --- | --- | --- | --- | --- | --- | --- | --- | --- | --- | --- |
| Spearman's rho | Substance use | Correlation Coefficient | -.067 | -.116* | .080 | .009 | -.007 | .122* | -.116* | -.153** |
|  |  | Sig. (2-tailed) | .238 | .041 | .157 | .875 | .900 | .031 | .041 | .007 |

\*\* . Correlation is significant at the 0.01 level (2-tailed).
\* . Correlation is significant at the 0.05 level (2-tailed).

### Multiple logistic regression analysis of significant variables

We used the stepwise logistic regression analysis method to investigate substance use predictors. The final model includes male gender, a recent family history of substance use, and family satisfaction as significant predictors as shown in Table 8. Adolescent boys were 4.23 times more likely to use substances than teenage girls (CI=1.68 – 10.64, p<0.05). Respondents with a family history of recent substance use were 2.68 times more likely to use substances than those who are not using substances with no family history of recent substance use (CI= 1.11-6.43, p<0.05). Respondents with family satisfaction were 0.27 times less likely to use substances than those who did not have family satisfaction (CI= 0.08-0.87, p<0.05).

**Table 8.** Multiple logistic regression for substance use.

| **Variables** | **B** | **SE** | **P-value** | **Odds Ratio** | **95% Confidence Interval** |
| --- | --- | --- | --- | --- | --- |
| Gender (Boy) | 1.44 | 0.47 | **0.002** | 4.23 | 1.68 – 10.64 |
| Family history of recent substance use | 0.99 | 0.45 | **0.028** | 2.68 | 1.11 – 6.43 |
| Family Flexibility | 0.25 | 0.77 | 0.746 | 1.28 | 0.285 – 5.76 |
| Family Chaotic | 0.24 | 0.59 | 0.687 | 1.27 | 0.40 – 3.99 |
| Family Communication | 0.41 | 0.69 | 0.555 | 1.50 | 0.39- 5.83 |
| Family Satisfaction | -1.32 | 0.60 | **0.028** | 0.27 | 0.08 – 0.87 |

B: beta; SE: standard error

## Discussion

Identifying the pattern of substance use among adolescents is crucial for delivering the best prevention, intervention, and harm reduction strategies. This study investigated the relationship between religiosity and family functioning with substance use and factors associated among Secondary school students in Northern Malaysia, a high-risk area.

The prevalence of substance use among adolescents is 9.6%. The prevalence is consistent with a study conducted in Egypt for polysubstance use [23]. This study found a lower prevalence than studies from other countries, including Timor-Leste, the United States, Norway, and Iran [24–27]. These disparities in findings could be attributed to the study’s data collection method (i.e. online survey method), sampling size, and respondent characteristics, while other research used high school and university students and a broader age range (age 16-40) as samples. Other causes may include cultural differences and social customs.

The present study shows cigarettes (4.8%) are the most often used substance, followed by alcohol (2.2%), stimulants (1.6%), inhalants (1.3%), and cannabis (1.3%). This finding is consistent with prior local studies that indicated current tobacco and alcohol usage among those in secondary school (15 to 18 years old) to be 6.6% and 2.4%, respectively [28]. De la Torre-Luque and colleagues discovered that tobacco was the most often used substance among adolescents in Indonesia, Laos, Malaysia, Myanmar, the Philippines, and Thailand [29]. However, the present study showed a lower percentage of tobacco and alcohol use compared to the NHMS 2022, which revealed 9.0% and 7.4% respectively. It also exhibited a lower prevalence when compared to other majority Muslim populations or ASEAN countries [30–32]. One systematic study found that nicotine product usage was a strong predictor of future cannabis use among adolescents [33].

This study revealed that a higher proportion of substance use were males (56.7%) and Malays ethnicity (99.0%). Even though most of the respondents are female, the study still shows male gender (16.3%) contributed a higher substance use compared to females (6.3%). The prevalence was consistent with the Department of Statistics Malaysia in 2020 revealing substance users are greater among the male gender, Bumiputeras, and highest education level secondary school. According to the National Health and Morbidity Survey (NHMS 2022), three out of every four drug users began using drugs before the age of 14. This finding is in keeping with the present study as we found that early adolescents (13-14 years old) were more likely to use substances. Whereas according to the World Health Organization(WHO) and other overseas studies [23,34] higher substance use among adolescents aged 14 – 17 years old. These disparities can be attributed to the generalizability of the studies, which included a broader range of adolescent ages than our study. Despite the country’s smoking prohibition for adolescents under the age of 18, many factors lead to ongoing smoking within this age group. The peer pressure and the desire to fit in with peers who smoke can encourage adolescents to begin and maintain smoking. Furthermore, easy access to tobacco products via older peers or family members can get beyond regulatory prohibitions. The portrayal of smoking in media and entertainment can further glamorize the habit, making it appear attractive and sophisticated. They can also purchase cigarettes from commercial sources according to research conducted at selected secondary schools in Peninsular Malaysia [35]. Adolescents may also be influenced by family views toward smoking, as parental or sibling smoking might normalize the activity. The gateway theory can explain why licit substances, such as tobacco and alcohol, are frequently used before more illegal substances. This hypothesis stated that drug use happens in stages, beginning with the use of licit substances like tobacco and alcohol and progressing to the use of illicit drugs.

Adolescent substance use can also be significantly influenced by family members who use substances. Our findings indicate that 16.8% of adolescents had a family history of recent substance use. This finding was consistent with other studies that identified familial environment as a social element in youth substance use [11,36–39]. We based our hypothesis on Albert Bandura’s social learning theory and Ajzen’s theory of planned behavior. Bandura hypothesized that any social learning environment contains both models and learners. In this social learning context, learners witness and imitate their models’ activities, either purposefully or unintentionally, which may explain why adolescents use substances more commonly when they have family members who use substances.

Adolescents from families with low incomes frequently endure increased stress and instability, which might lead them to seek joy or escape through substance use. Parents in low-income families may have to work several jobs or long hours, which results in less supervision and engagement with their children, making adolescents more exposed to negative peer pressures and risky behaviors. Inadequate financial resources might also limit access to community activities and support networks that promote healthy development and substance use prevention. Several studies have found a link between low-income families and adolescent substance use [40–42]. This is consistent with the current study, which indicated that most of the adolescents who use substances come from low-income families. A few studies have shown that adolescents from lower parental education levels are more likely to use substances [11,40,41]. This is in keeping with the present study that most adolescents who use substances also have parents with a level of secondary school education or can be considered among low education level. Their parents may face various socio-economic challenges, including lower income, and be less aware of the hazards and consequences of substance use, resulting in less effective communication and monitoring for drug and alcohol prevention. Economic issues related to poorer educational attainment can cause increased stress in the home, producing settings in which adolescents may turn to substances as a coping mechanism.

Overall, the respondents in our study have balanced family functioning which balanced scales showed connected in cohesion level with flexible in flexibility level. In addition, the unbalanced scales show low levels for disengaged and enmeshed, very low levels for chaotic, and moderate levels for rigid [42]. Family functioning in terms of balanced flexibility, family satisfaction, and communication shows a negative correlation with substance use indicating that high family flexibility, family communication, and family satisfaction, as assessed by the FACES-IV, play protective roles in preventing substance use among adolescents due to their contributions to a supportive and adaptive family environment. High family flexibility refers to a family’s ability to adjust to changes, handle stress, and reorganize roles and responsibilities. This adaptability contributes to a stable yet dynamic environment in which adolescents feel supported and understood, lowering the likelihood of them turning to substances as a method of coping with stress or uncertainty. Effective communication within the family is essential for building trust, understanding, and emotional support. When family members talk openly and honestly, adolescents are more likely to share their concerns, seek advice, and feel valued. This open communication can aid in detecting early signs of distress or harmful behaviors, allowing for prompt interventions. High family satisfaction reflects a positive and rewarding family experience, as evidenced by strong emotional relationships, mutual support, and a sense of happiness within the family. Adolescents who are happy with their family relationships are more likely to internalize positive family values and norms. These 3 aspects will discourage the adolescent from seeking comfort or validation through substance use. The present study also shows there is a significant positive correlation between a chaotic family and substance use implying that an adolescent who had a more chaotic family would increase substance use. Our findings are consistent with previous research highlighting the importance of a healthy family dynamic and the parent-adolescent relationship in reflecting whole-family functioning, which can be protective of adolescent substance use [43–48].

Previous research has found that people with low religiosity scores use more substances. Previous studies also show a significant association between substance use and religiosity, with strong religiosity being related to a reduced rate of substance use due to a conservative attitude against substance use and improved psychological well-being [9,48–50]. However, our findings reveal that there is no association between religiosity and substance use, even though respondents had higher religiosity scores for the knowledge and practice component. While religiosity often promotes beliefs and actions that hinder substance use, its impact can be diminished by other relevant factors in an adolescent’s life despite being born Muslim. Peer pressure and the desire for social approval can sometimes override religious principles, particularly during a developmental stage marked by experimentation and the formation of identity. Furthermore, the level and intensity of religious participation can vary; for some adolescents, religious practices may be more superficial or culturally expected than profoundly internalized, limiting their influence on behavior. Adolescents from strongly religious families may face strictness, which can lead to rebellious and secretive behaviors, including substance use, as a form of protest. These complicated interactions demonstrate that, while religiosity can be a protective factor, it is not an absolute barrier to substance use in adolescents.

Our final analysis using logistic regression reveals male gender, recent family history of substance use, and family satisfaction as significant predictors of substance use among adolescents. The present study found that male adolescents are four times more likely than female adolescents to use substances, which is consistent with previous studies indicating that males use substances more than females [23,36,37,39,40,51]. Our study shows the likelihood of substance use among adolescents was 2.68 times (CI: 1.11 - 6.43) higher among adolescent boys from substance-using families. This finding is consistent with previous studies showing that adolescents are more likely to use substances if their family members use substances [52,53].

Family satisfaction is a protective factor, making adolescents 0.27 times (CI: 0.08-0.87) less likely to use substances. It is one of the domains of FACES-IV that assesses how satisfied adolescents are with their relationships with other family members and family dynamics. This scale assesses communication, emotional bonding, support, problem-solving skills, and adaptability to change and stress. Higher scores on this measure suggest higher contentment and perceived effectiveness in family functioning, whereas lower scores may indicate areas where family members are detached or unsatisfied. Previous research has also highlighted the importance of family satisfaction as a whole family functioning be a protective factor against substance use among adolescents [54].

In summary only male gender, family history of substance use, and family functioning, specifically family satisfaction were shown as significant predictors of substance use among adolescents. Religiosity was not significant with substance use.

### Study limitations and recommendations

Our study has a few limitations, and the findings must be carefully evaluated. First, the study design is cross-sectional, hence only allowing the determination of the association between dependent and independent variables but not causal relationships. Second, the substance use status of adolescents was obtained by self-reporting without any other objective validation such as urine drug toxicology. Some respondents may remain apprehensive, and concerned about how the information provided to the researchers may be used due to possible legal implications. Third, the study was limited to a selected secondary school in the Northern area of Malaysia and included only students aged 13 to 16 which reduces the study’s capacity and generalization of the findings. This study focuses only on Muslim adolescents, and it would be beneficial if similar studies included a variety of religions and ethnicities to reflect the population of adolescents in Malaysia since Malaysia is a multiracial and multicultural country.

Despite certain limitations, these findings possess significant implications for future interventions to address the problem, thus effective interventions should be multifaceted.

Parents play an important role in fostering coping skills and drive for their children’s achievement. Parents should be offered interaction and supervision skills training, as well as family therapy to improve family relations and satisfaction. Policymakers and schools should collaborate to provide materials and workshops to assist parents in better understanding adolescent development and good parenting methods. Such support can help parents identify signs of distress in their children and provide appropriate support to alleviate the problem.

Schools need to implement comprehensive substance use prevention programs, integrate mental health services, and promote extracurricular activities that engage students in positive ways. Regular and continuous psychoeducation for adolescents about the implications or adverse consequences of substance use, which began with smoking, is critical to preventing future use of illicit drugs. This purpose should be incorporated into the school module and emphasized by potent school policy. Much attention must be taken to prevent any “soft” substance commencing (tobacco and alcohol), particularly at a young age. Tobacco, alcohol, and illegal drug control programs should prioritize reducing youth access to these substances. For at-risk students, the school has to offer prompt interventions that are characterized by poor coping skills and diminishing academic performance. These initiatives may include counseling services, academic support programs, and referrals to external resources. Recognizing and addressing mental and emotional challenges at an early stage may help adolescents avoid resorting to drugs as a coping method. Adolescents will benefit from having access to counseling and mentorship programs, which can help them develop resilience and appropriate coping skills.

Public health initiatives should be established to educate adolescents about the risks of drug use. These efforts should address adolescents and their parents to underline the necessity of maintaining good academic achievement as well as the potential harmful effects of substance use on academic success and future opportunities.

Policymakers have to set up mechanisms for monitoring and evaluating the efficacy of coping skills interventions and therapies offered in schools. Regular assessments can help identify areas for development and ensure that resources are spent efficiently on programs that yield positive results. Such interventions may enhance the coping skills and academic performance of vulnerable adolescents. Enforcing strict regulations on substance availability to minors, increasing funding for prevention and treatment programs, and launching public awareness campaigns are essential as well. These interventions can collectively address the root causes and protective factors of adolescent substance use, creating a supportive environment that reduces risk and promotes healthy development.

Relevant organizations and agencies must also take a more proactive approach to assisting and monitoring adolescents to safeguard their well-being and prevent additional substance use disorders or dependence.

## Conclusion

Our study involved adolescents from selected schools from hotspot substance areas which can provide insights that are directly applicable to areas where intervention is most needed.

In conclusion, these findings can be used to reinforce existing policy initiatives for adolescents to establish a drug-free environment. Emphasizing the importance of parents and family members in recognizing early signs of distress, better communication, and support may help at-risk adolescents develop better ways to cope despite their substance use. The significant consequences of the study include heightened awareness, prompt identification, and intervention for adolescents and parents through both universal school-based and tailored programs. Preventing substance use in early adolescence requires a collaborative effort among parents, schools, public health professionals, policymakers, and other stakeholders because of the multifaceted nature of the issue. By working together, these groups can create a consistent and reinforced message, address the social determinants of health, and build a resilient community that supports the healthy development of adolescents, ultimately reducing the incidence of substance use and its associated consequences. Nonetheless, the data were self-reported, and focusing on school-age adolescents may exclude high-risk adolescents who have dropped out of school.

## Data Availability

All relevant data are within the manuscript.

## Author contributions

Conceptualization: Noor Adnin binti Ab Aziz, Suzaily Wahab, Rosnah binti Sutan

Data curation: Rosnah binti Sutan, Muhammad Adib Baharom, Amirul Danial Azmi

Formal analysis: Rosnah binti Sutan, Siti Azirah binti Asmai

Funding acquisition: Suzaily Wahab

Methodology: Noor Adnin binti Ab Aziz, Suzaily Wahab, Rosnah binti Sutan

Project administration: Noor Adnin binti Ab Aziz, Suzaily Wahab

Supervision: Suzaily Wahab.

Writing – original draft: Noor Adnin binti Ab Aziz

Writing – review & editing: Suzaily Wahab, Rosnah binti Sutan, Muhammad Adib Baharom

## Acknowledgment

The authors would like to express their gratitude to all students who were involved in this study. We would like to thank the Director General of Health Malaysia for his permission to publish this article. We acknowledge the Medical Faculty of the University Kebangsaan Malaysia who support this publication.

